# Interoceptive disruption in functional neurological disorder: a multimodal brain imaging study

**DOI:** 10.1101/2024.03.13.24303546

**Authors:** Petr Sojka, Tereza Serranová, Sahib S. Khalsa, David L. Perez, Ibai Diez

## Abstract

This multimodal brain imaging study investigated functional MRI (fMRI) neural processing of cardiac interoceptive signals in 38 patients with functional neurological disorder (FND) compared to 38 healthy controls (HCs). Additionally, we characterized how brain fMRI responses during heartbeat counting (interoception) vs. tone counting (exteroception) or rest related to grey matter volume, interoceptive awareness, and psychopathology scores. For both interoception vs. rest and interoception vs. exteroception contrasts, principal component analyses showed that principal component 1 (PC1) as derived from all study participants was comprised primarily of salience, ventral attention and sensorimotor network co-activations, along with default mode and visual processing network co-deactivations. Compared to HCs, patients with FND showed reduced contribution to these PC1 co-activation/co-deactivations patterns in both interoception vs. exteroception and interoception vs. rest contrasts; only the interoception vs. exteroception between-group fMRI findings held adjusting for depression/anxiety scores, antidepressant use and FND subtype. For the interoception vs. rest contrast, increasingly negative PC1 contribution scores positively correlated with decreased cingulate gyrus volumes and increased psychopathology scores. This multimodal brain imaging study underscores a role for salience and default-mode networks in the pathophysiology of FND, and sets the stage for comprehensive research efforts further contextualizing the mechanistic importance of altered interoception in patients with FND.

## 1. Introduction

Functional neurological disorder (FND) is a condition at the intersection of neurology and psychiatry manifesting in convulsions, abnormal movements, sensory deficits, dizziness and/or cognitive symptoms (Hallett et al., 2022). Functional neurological symptoms are not primarily driven by a discrete structural lesion, but rather reflect abnormal activations within and across several distributed brain networks (Baizabal-Carvallo et al., 2019; Hallett et al., 2022. Core FND and non-motor symptoms (e.g., fatigue, pain, dissociation, depression and anxiety) lead to high disability and impaired quality of life comparable with those observed in other major neuropsychiatric conditions (Campbell et al., 2022; Forejtová et al., 2023). Traditionally, emphasis has been placed on psychophysiological factors such as emotion dysregulation and adverse life experiences as predisposing vulnerabilities and/or acute precipitants in patients with FND (Drane et al., 2020). However, bodily-related events such as physical injury, acute illnesses, and vaccinations can also precipitate or trigger FND (Butler et al., 2021a; Stone et al., 2009). Interestingly, affectively-valenced psychological experiences and physical events both engage the interoceptive nervous system (Carvalho & Damasio, 2021), with interoception recognized as a mechanistically important construct in updated pathophysiological models of FND (Baizabal-Carvallo et al., 2019).

Interoception is the process by which the nervous system senses, integrates, and interprets signals originating from within the body (Khalsa et al., 2018). In FND, abnormal top-down predictions about internal bodily states and/or aberrant bottom-up sensory information may promote the brain to inaccurately model the body (Edwards et al., 2012; Koreki et al., 2022; Lin et al., 2020). Potential disruptions in interoception have been proposed to contribute to the development and/or maintenance of FND through impairments in allostasis (i.e., the active process of forecasting the energetic needs of the body by modeling the body in the world) (Jungilligens et al., 2022). Additionally, it was recently theorized that the brain may in some instances inefficiently categorize physical signals across the spectrum of valence and arousal (e.g., panic attack without panic) (Jungilligens et al., 2022). A failure to adaptively model interoceptive signals may also be a transdiagnostic vulnerability in mechanistically-related or comorbid neuropsychiatric conditions such as functional somatic disorders, (e.g., fibromyalgia), somatic symptom disorder, depression, and anxiety (Khalsa et al., 2018; Paulus et al., 2019).

Thus, it is notable that limited research has investigated interoceptive processing in FND to date. Mismatches between objective and subjective symptom reports or between measured physiological responses and emotional experiences identified in FND have been interpreted as support for abnormal interoceptive inference (Adewusi et al., 2021). Furthermore, a few studies used heartbeat counting tasks to experimentally probe interoceptive accuracy in FND cohorts. However, this research has yielded mixed findings – with reports of both lower interoceptive accuracy (Koreki et al., 2020; Ricciardi et al., 2021; Williams et al., 2021) or no difference compared to healthy controls (HCs) (Jungilligens et al., 2020; Koreki et al., 2023; Millman et al., 2023; Sojka et al., 2021); these differences may relate to heterogeneity in the neural mechanisms for FND across populations and/or to methodological considerations (e.g., small sample sizes, differences in heartbeat detection paradigms, or poor resting cardiac interoceptive awareness in the general population leading to floor effects). Of note, the tendency to overestimate interoceptive heartbeat counting accuracy (labeled as interoceptive trait prediction error, ITPE) in patients with functional seizures positively correlated with seizure frequency (Koreki et al., 2020), highlighting the mechanistic relevance of individual differences in interoceptive accuracy. Even when interoceptive accuracy deficits are not appreciated, there may be distinct (and less efficient) neural circuit alterations mediating interoception that could contribute to the pathophysiology of FND (Eikommos et al., 2023).

Neurobiologically, interoception is supported by portions of the salience, default mode and ventral attention networks (Kleckner et al., 2017). Systems neuroscience research proposes that the default mode network initiates predictions, while the salience network estimates the importance of afferent signals and contributes to prediction error learning (Kleckner et al., 2017). The temporoparietal junction (TPJ), implicated in impaired action-authorship perceptions across several FND studies, is at the intersection of the salience, default mode and ventral attention networks (Drane et al., 2020; Yeo et al., 2011). In FND, cingulo-insular neuroimaging profiles have correlated with symptom severity (Diez et al., 2019; Li et al., 2014; Perez et al., 2021) and indices of disease risk (Maurer et al, 2016). A recent multicenter study identified cingulo-insular, inferior parietal lobule, primary sensorimotor, and hippocampal resting-state functional connectivity patterns as the most discriminant features distinguishing FND vs. HCs in a machine learning classification (Weber et al., 2022). In task-related emotion processing and motor fMRI studies in FND cohorts, altered visceromotor activations (i.e., insula, cingulate gyrus, ventromedial prefrontal cortex (vmPFC)) were also reported (Aybek et al., 2015; Goodman et al., 2022; Voon et al., 2011). However, only one neuroimaging study has investigated correlates of interoception in FND to date, identifying that decreased white matter integrity from the insula and TPJ related to reduced interoceptive accuracy in a mixed FND cohort (Sojka et al., 2021).

This confluence of evidence suggests possible network disruptions related to interoceptive processing in FND. We therefore hypothesized that patients with FND would exhibit abnormal activations in major interoceptive hubs (e.g., anterior insula, cingulate gyrus, vmPFC) during the processing of internal bodily signals compared to HCs. To test this hypothesis, 38 patients with mixed FND and 38 HCs performed an fMRI heartbeat tracking task (HTT), along with exteroceptive and rest conditions. Compared to comparison conditions, we hypothesized that patients with FND vs. HCs would demonstrate altered salience, default mode and ventral attention network activations during cardiac interoceptive processing. We also hypothesized that functional activations in these networks would relate to individual differences in interoceptive accuracy, ITPE and symptoms of dissociation, depression and anxiety in patients with FND during interoceptive processing. To reduce the dimensionality of the fMRI data and thus limit multiple comparison testing concerns, principal component analysis (PCA) was used for all fMRI findings (Mwangi et al., 2014). In parallel, voxel-based morphometry (VBM) was used to contextualize how individual differences in grey matter (GM) volumetric profiles related to fMRI activation.

## 2. Methods

### 2.1. Participants and Questionnaires

The study was approved by the Masaryk University and St. Anne’s University Hospital ethics committees and all participants provided written informed consent. Thirty-eight adults with FND (32 women, 6 men; age=34.7±13.2; average illness duration=4.3L±L2.8) meeting “rule in” diagnostic criteria for functional seizures (FND-seiz; F44.5) or functional movement disorder (FND-movt; F44.4) were recruited consecutively from the St. Anne’s University Hospital in Brno between April 2016 and June 2019. All patients had symptoms for over 2 years. This mixed FND cohort was comprised of 21 video-electroencephalography documented FND-seiz (19 women, 2 men; age=23.0L±L2.8; *n*=6, major motor; *n*=8, minor motor; *n*=7, atonic) and 17 clinically-established FND-movt (12 women, 5 men; age=44.0L±L12.7; n=6, tremor; n=3, gait disorder; n=2, dystonia; n=2, tics/jerks; n=3, mixed; n=1, weakness) patients. We used a transdiagnostic approach across the spectrum of FND-seiz and FND-movt given that mixed symptoms are common, and individuals that present with one clinical phenotype can develop new/distinct functional neurological symptoms over the natural history of their illness (Butler et al., 2021b; Matin et al, 2017; McKenzie et al., 2011). Thirty-eight HCs (32 women, 6 men; age=34.8±14.1) were recruited from the community through local advertisements. Demographic matching variables included self-reported age, sex and years of education. Exclusion criteria for all participants included age <18 years old, known MRI abnormality, intellectual disability, other major neurological/medical conditions (i.e., epilepsy or Parkinson’s disease), and psychotic/bipolar/substance use disorders. All participants were instructed not to take as needed CNS-acting medications two days prior to scanning and to abstain from caffeine, smoking and alcohol on the day of scanning. Participants were monetarily compensated for their participation. Note, these FND and HC cohorts have been previously published in the context of a diffusion tensor imaging study (Sojka et al., 2021).

Prior to scanning, participants completed the Beck Depression Inventory-II (BDI-II)(Beck et al., 2011), Spielberger State–Trait Anxiety Inventory (STAI)(Spielberger, 2012), Dissociation Experiences Scale-II (DES) (Carlson & Putnam, 2022) and Body Perception Questionnaire (BPQ) (Cabrera et al., 2018). For the BPQ, only the awareness (BPQ-aware) subscale was used, providing a trait measure of subjective beliefs about sensitivity to internal sensations, irrespective of their objective interoceptive performance (Garfinkel et al., 2016).

### 2.2. Paradigm

Cardiac interoceptive awareness was assessed with a heartbeat tracking task (HTT) that has been previously utilized to test interoceptive abilities in clinical populations including those with FND (Garfinkel et al., 2016; Koreki et al., 2020). During fMRI recording, participants were asked to count their heartbeats or acoustic tones perceived during 12 pseudo-randomly ordered time windows of varying length (15, 18, 21, 24, 27 and 30 seconds) and after each trial to report the number of heartbeats/tones counted (See **Figure 1** for task design). Twelve resting-state trials of the same length were interspersed between the experimental trials. Participants were asked to start counting heartbeats or tones when a heart/note pictogram appeared on a screen and to stop counting when a response window appeared. Participants were instructed to count only the heartbeats they confidently felt and asked to not guesstimate heartbeat counts.

**Figure 1.**
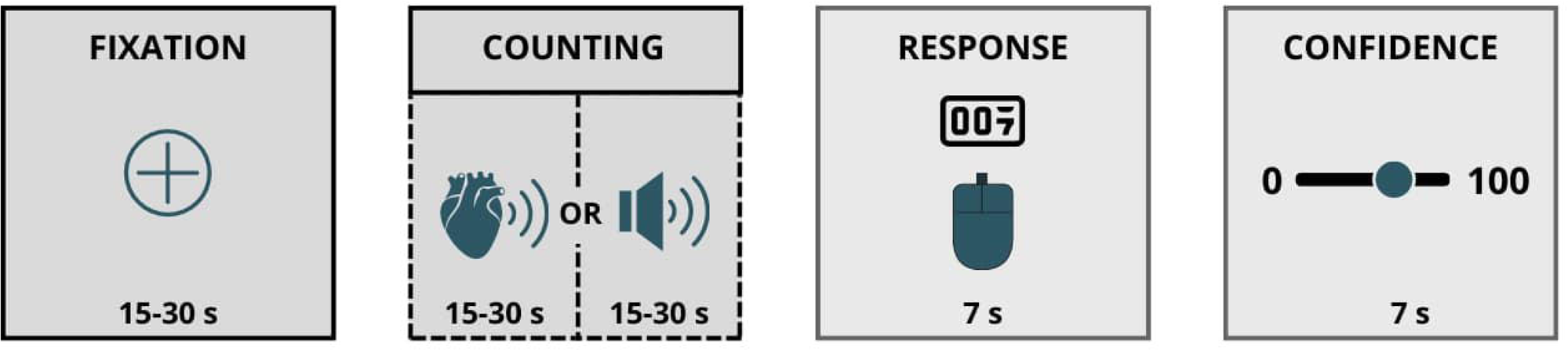
fMRI task design. Participants were asked to count their own heartbeats or auditory tones during 12 pseudo-randomly ordered time windows of varying length (15, 18, 21, 24, 27 and 30 seconds), report the number of heartbeats/tones counted, and indicate their confidence in their estimations. Twelve resting-state trials with the same length were interspersed in between the experimental trials.

Responses were made on a 2-button box, and the task was presented with E-Prime (Psychology Software Tools Inc, Pittsburgh) during fMRI recording. Participants’ heartbeats were monitored via an electrocardiogram (ECG) monitor (BrainVision BrainAmp MR) with three electrodes attached to the chest. To approximate the same task difficulty for both the interoceptive and the exteroceptive conditions, a staircase procedure was used prior to the experimental trials to set the intensity of the acoustic stimuli (pitch=440Hz, duration=750ms, repetition=1000ms) at a “just noticeable threshold” while the scanner was running. To ensure basic familiarity with the task prior to in-scanner performance, all participants first practiced three rounds of heartbeat tracking outside the scanner.

### 2.3. ECG Preprocessing

Due to induced artifacts from the fMRI sequence, the ECG data required preprocessing to extract R-peaks from the ECQ recordings during the task. R-peaks were extracted from the ECG in BrainVision Analyzer 2.1. (Brain Products) after MRI artifact correction and applying an IIR Butterworth low-pass filter. Semi-automated procedures were implemented, with R-peaks first automatically labeled in BrainVision analyzer and subsequently visually inspected; any missed or incorrectly labeled R-peaks were manually edited by graduate students blinded to group identity. As a final inspection of the raw data, author P.S. visually inspected all tracings to ensure accuracy of R-peak selections — which were readily apparent across all participants with usable data.

### 2.4. Behavioral Data Analyses

A heartbeat counting accuracy score (HACC) reflecting interoceptive accuracy was calculated according to the following transformation:

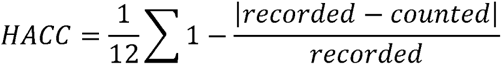

Scores vary between 0 – 1, with higher scores indicating better interoceptive accuracy (task performance). Interoceptive trait prediction error (ITPE) was calculated as the difference between subjective interoceptive sensitivity (BPQ-aware) and interoceptive accuracy (Garfinkel et al., 2016). The larger and more positive the value of ITPE, the greater the interoceptive error at the trait level.

Independent t-tests were used to calculate group differences in BDI-II, STAI-total, DES, BPQ-aware, interoceptive accuracy and ITPE scores. Where the Levene test for equality of variances was violated, df, t-values and significance values were adjusted using the Welch t-test. False discovery rate corrected significance values for multiple statistical testing concerns. All reported significance levels are two-tailed.

### 2.5. Scanning Parameters

Brain MRI scans were acquired with a Siemens 3 Tesla Magnetom Prisma scanner at the Central European Institute of Technology in Brno. First, a high-resolution anatomical T1-weighted magnetization prepared rapid gradient-echo (MPRAGE) sequence was acquired with the following parameters: 1mm isotropic voxels, 240 sagittal slices, slice thickness 1.00mm, acquisition matrix size=252×224, repetition time (TR)=2,300ms, echo time (TE)=2.33m, and flip angle (FA)=8. Subsequently, a whole-brain blood oxygenation level dependent (BOLD) functional scan was acquired, while the participant performed an interoception-exteroception-rest task, using multiband acquisition with the following parameters: TR=642ms, TE=35.0ms, FA=47°, voxel size 3.3×3.3×3.5mm, 40 axial slices, field of view 210×210 mm.

### 2.6. MRI Preprocessing

MRI data was preprocessed using FMRIB Software Library v5.0.7 (FSL) and MATLAB R2019b. The preprocessing of the anatomical T1 image included: reorientation to right-posterior-inferior (RPI) and alignment to anterior and posterior commissures; skull stripping; GM, white matter and cerebrospinal fluid segmentation; and calculation of non-linear transformation between T1 and 2mm resolution MNI152 standard template. The preprocessing of the functional MRI included: slice timing correction; reorientation to RPI; motion correction (rigid-body realignment of functional volumes within runs); computation of the transformation between individual skull-stripped T1 and mean functional images using linear boundary-based registration(https://fsl.fmrib.ox.ac.uk/fsl/fslwiki/FLIRT_BBR); intensity normalization; non-linear transformation of data to MNI space, concatenating the transformation from functional to structural and from structural to 3mm MNI standard space; spatial smoothing with an isotropic Gaussian kernel of 6mm FWHM; and a high-pass filtering of 0.015 Hz to reduce noise and low-frequency drift.

### 2.7. fMRI Multivariate Data Analyses

Task fMRI analysis was performed using the general linear model (GLM) approach in FEAT (FMRIB’s Expert Analysis Tool) to derive individual subject contrast maps. Specifically, a first-level GLM was used to model the experimental design at voxel-level using 3 regressors: interoception, exteroception and rest blocks. The 3 regressors were convolved with a double Gamma canonical hemodynamic response function. Temporal derivatives of these three regressors were also added to compensate for variability in the hemodynamic response function. Additionally, extra regressors were included to control for different sources of noise: mean signal of white matter, mean signal of CSF and 6 head motion parameters. Individual subject contrast maps-of-interest included: interoception vs. rest and interoception vs. exteroception.

PCA was used to reduce the voxel-level dimensionality of the findings by separating out the interoception vs. rest and interoception vs. exteroception maps into a set of covarying activation/deactivation patterns that explained most of the variance (Mwangi, Tian & Soares, 2014). First, the contribution of age, sex, and BMI to the interoception vs. rest and interoception vs. exteroception maps were computed using a GLM and removed from the data. Then, for each contrast, PCA was applied to the images of all FND and HCs. The PCA returns a set of principal components (PCs) - spatial maps - with weight assigned to each voxel and scores representing how much each subject contributes to the PC. These PC scores were used to compute between-group differences and within FND correlations. To derive the component spatial map, we used a GLM and computed the association between each component scores with interoception vs. rest and interoception vs. exteroception maps. Only voxels with a p-value < 0.05 surviving to multiple comparisons are displayed. Whole-brain correction for multiple comparisons was computed using Monte Carlo simulation with 10,000 iterations to estimate the probability of false positive clusters with a two-tailed p-valueL<L0.05 (3dClustSim, afni.nimh.nih.gov).

For statistically significant between-group fMRI results, separate *post-hoc* analyses attempted to adjust for: 1) depression (BDI-II) and anxiety (STAI-total) scores; 2) antidepressant medication use (yes/no); and 3) FND-seiz subtype (yes/no).

In addition to between-group analyses, we also tested relationships between individual-subject contributions to a given component and the following 5 measures: HACC, ITPE, DES, BDI-II and STAI-total scores. In these within-group correlational analyses, false discovery rate corrected significance values for multiple statistical testing concerns.

### 2.8. Voxel-based morphometry

To study how voxel-based GM volumetric profiles related to interoception vs. rest and interoception vs. exteroception PC scores at the individual-subject level, FSL VBM was used (Douaud et al., 2007), an optimized VBM protocol (Good et al., 2001) carried out with FSLtools (Smith et al., 2004). T1-weighted images were brain-extracted and GM was segmented before registering to the MNI 152 standard space using non-linear registration. The resulting images were averaged and flipped along the x-axis to create a left-right symmetric, study-specific GM template. All native GM images were non-linearly registered to the created study-specific template and “modulated” to correct for local expansion (or contraction) due to the non-linear component of the spatial transformation. The modulated GM images were then smoothed with an isotropic Gaussian kernel with a sigma of 3.

A GLM was used to study the association between fMRI PC scores (specifically PC1, as this component was the only statistically significant component showing between-group differences: see results) and voxel-level GM volumes adjusting for total intracranial volume, age, sex, and BMI. Monte Carlo simulation was used to correct for multiple comparisons with 10,000 iterations to estimate the probability of false positive clusters with a two-tailed p valueL<L0.05 (3dClustSim, afni.nimh.nih.gov).

### 2.9. Data Availability

De-identified data related to study findings and not published within this article will be made available by request from any qualified investigator following local IRB approval.

## 3. Results

### 3.1. Behavioral Findings

See Sojka et al., 2021 for previously reported behavioral/psychometric findings from this same cohort. In brief, patients with FND and HCs were matched for key demographic variables with no significant differences in age, sex, BMI, and years of education. However, patients with FND reported elevated dissociation, depression and anxiety scores compared to HCs. BPQ-awareness scores were not statistically different between groups (See **Table 1**). Due to noise and artifact levels, ECG data from 5 FND patients and 4 HCs were excluded. Thus, for the HACC and ITPE within-group analyses, 33 FND patients (age=34.7L±L13.2; M=6; F=27; FND-seiz=18; FND-movt=15; Body Mass Index (BMI)=24.3L±L5.4) and 34 HCs (age=34.8L±L14.1; M=6; F=28; BMI=23.5L±L4.0) were used. In those with available ECG data, there were no statistically significant group differences in HTT scores (FND: 0.63L±L0.26; HCs: 0.70L±L0.21; *p*=0.41) when controlling for the effect of BMI (and there were also no significant FND-movt vs. FND-seiz vs. HCs subgroup effects in HTT scores). ITPE values did not significantly differ between the two groups (FND: 0.39±1.46; HCs: - 0.29±1.43; *p*=0.20).

**Table 1.** Demographic and clinical information of the functional neurological disorder (FND) and healthy control (HC) cohorts. Note, 11 patients with FND were on selective serotonin reuptake inhibitors and/or serotonin norepinephrine reuptake inhibitors. BMI: Body Mass Index; BDI-II: Beck Depression Inventory-II; STAI: Spielberger State Trait Anxiety Inventory; DES: Dissociation Experiences Scale; BPQ: Body Perception Questionnaire; y: years.

| Variable | Mean (SD) |  | t-value (df) | p-value |
| --- | --- | --- | --- | --- |
|  | FND (n = 38) | HCS (n = 38) |  |  |
| Age, y | 34.7 (13.2) | 34.8 (14.1) | 0.02 (74) | 0.98 |
| Education, y | 12.0 (2.3) | 12.3 (2.6) | 0.53 (74) | 0.61 |
| BMI | 24.3 (5.5) | 23.5 (4.1) | 0.76 (74) | 0.45 |
| BDI-II | 20.1 (11.5) | 11.0 (6.9) | 4.22 (74) | 0.006 |
| STAI-total | 82.2 (17.2) | 71.1 (17.1) | 2.73 (72) | 0.008 |
| DES | 26.5 (11.2) | 18.6 (7.5) | 3.44 (72) | 0.001 |
| BPQ-awareness | 112.6(28.1) | 100.4 (26.8) | 1.40 (74) | 0.11 |

### 3.2. Between-Group PCA Findings

#### 3.2.1. Interoception vs. Rest

Four PCs cumulatively had the most impact on explained variance in the data for the interoception vs. rest contrast (22% variance, See **Figure 2A**). Only contributions to PC1 (10% variance) showed between-group differences, with patients with FND contributing significantly less to this component than HCs (p=0.006; z=-2.71; see **Figure 2B**); these between-group findings held in *post-hoc* analyses adjusting for FND subtype, but did not hold adjusting for depression/anxiety scores or antidepressant use. This first PC showed significant synchronous BOLD co-activations in regions that are part of the salience, ventral attention and sensorimotor networks, including large clusters within the bilateral anterior insula, inferior parietal lobule/TPJ, paracentral lobule, and middle cingulate gyri. Conversely, this component exhibited significant synchronous BOLD co-deactivations in regions that are part of the default mode and visual processing networks, including large clusters in bilateral ventromedial prefrontal, occipital, precuneus and posterior cingulate brain areas. Co-deactivations were also observed in bilateral superior temporal gyri, the periaqueductal grey and hypothalamus.

**Figure 2.**
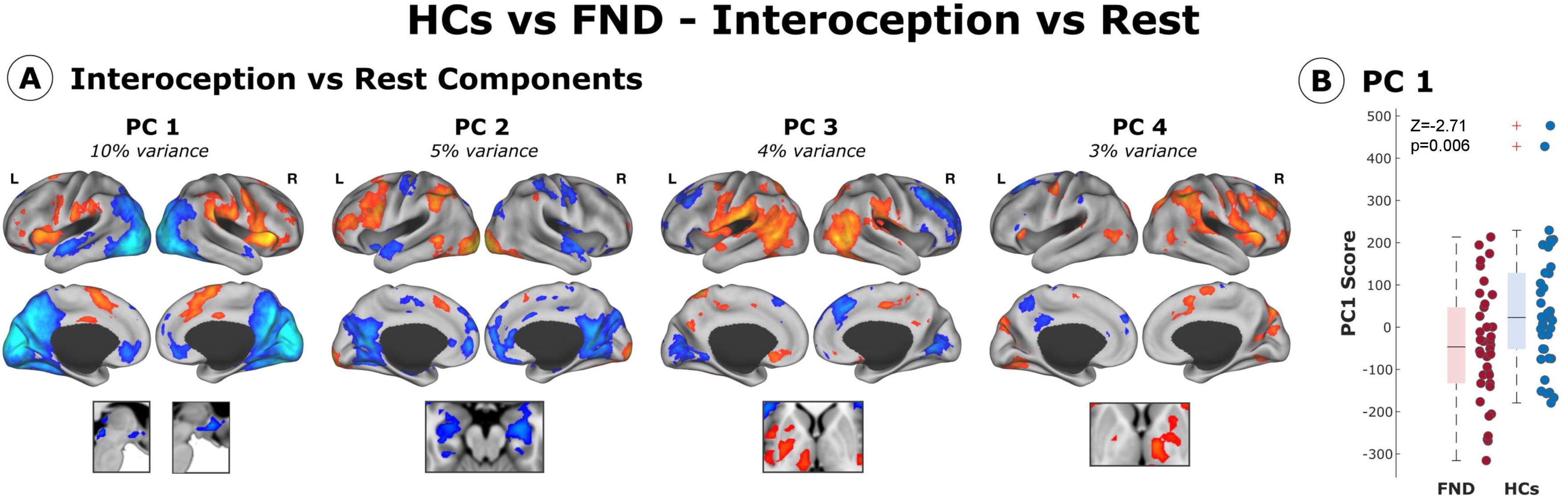
Principal components representing task-related variance in interoception > rest contrast. **A.** Four principal components (PCs) cumulatively explained most of the total variance in the data for the interoception vs. rest contrast. **B.** Only contributions to the PC1 showed between-group differences, with FND patients contributing significantly less to this component than HCs; these between-group differences remained statistically significant adjusting for FND subtype, but did not remain significant adjusting for depression/anxiety scores nor antidepressant medication use. PC1 was characterized by significant BOLD co-activations in key brain regions belonging to the salience, ventral attention, and sensorimotor networks. These included the bilateral anterior insula, inferior parietal lobule/TPJ, paracentral lobule, and middle cingulate gyri. Conversely, this component exhibited significant BOLD co-deactivations in regions that are part of the default mode and visual processing networks, including clusters in bilateral ventromedial prefrontal, occipital, precuneus and posterior cingulate brain areas. Co-deactivations were also observed in bilateral superior temporal gyri, the periaqueductal grey and hypothalamus. Note, removal of two potential outliers in the healthy control (HC) sample (identified with a *), did not impact the statistical significance of the between-group findings (z=-2.31; p=0.02).

#### 3.2.2. Interoception vs. Exteroception

Five PCs cumulatively had the most impact on explained variance in the data for the interoception vs. exteroception contrast (28% variance, See **Figure 3A**). Again, only contributions to PC1 (10% variance) showed between-group differences, with patients with FND contributing significantly less to this component than HCs (p=0.002; z=-3.05; see **Figure 3B**). In separate *post-hoc* analyses, these between-group findings held adjusting for depression/anxiety scores, antidepressant use and FND subtype. This first PC showed significant synchronous BOLD co-activations in regions that are part of the salience, ventral attention, central executive, and sensorimotor networks, including large clusters within the bilateral anterior insula, inferior parietal lobule/TPJ, paracentral lobule, middle cingulate/dorsal anterior cingulate gyri and middle frontal gyri, as well as the right inferior frontal gyrus. Conversely, this component exhibited significant synchronous BOLD co-deactivations in regions that are part of the default mode and visual processing networks, including large clusters in bilateral ventromedial prefrontal, occipital, precuneus and posterior cingulate brain areas. Co-deactivations were also observed in bilateral anterior temporal cortices, right hippocampus, right amygdala, and the periaqueductal grey.

**Figure 3.**
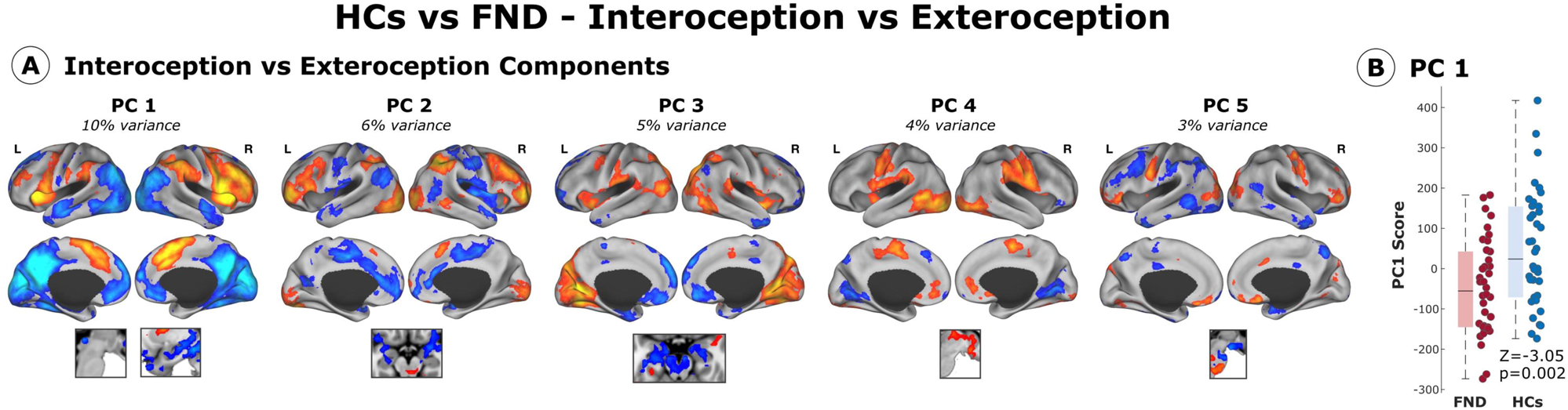
Principal components representing task-related variance in interoception > exteroception contrast. **A.** Five principal components (PCs) cumulatively explained most of the total variance in the data for the interoception vs. rest contrast. **B.** Only contributions to PC1 showed between-group differences, with patients with FND contributing significantly less to this component than HCs; these between-group findings remained statistically significant adjusting separately for 1) anxiety/depression scores, 2) FND subtype, and 3) antidepressant medication use. PC1 exhibited significant BOLD co-activations across salience, ventral attention, central executive, and sensorimotor networks. Notably, large clusters were observed in the bilateral anterior insula, inferior parietal lobule/TPJ, paracentral lobule, middle cingulate/dorsal anterior cingulate gyri, middle frontal gyri, and the right inferior frontal gyrus. Conversely, this component showed significant BOLD co-deactivations in default mode and visual processing networks, including large clusters within bilateral ventromedial prefrontal, occipital, precuneus, and posterior cingulate regions. Additional co-deactivations were noted in bilateral anterior temporal cortices, the right hippocampus, amygdala, and the periaqueductal grey.

### 3.3. Associations Between Differentiating Principal Component Scores and Grey Matter Volumes

In patients with FND, relative reductions in bilateral left > right GM volumes in the ventral and dorsal prefrontal cortices – including the rostral and dorsal anterior cingulate cortex – correlated with reduced PC1 contribution scores for the interoception vs. rest contrast (see **Figure 4**). In patients with FND, individual differences in GM volume bilateral dorsomedial prefrontal, right posterior temporal and left superior parietal cortices showed statistically significant associations with loading score contributions in PC1 for the interoception vs. exteroception contrast (see **Supplementary Figure 1**).

**Figure 4.**
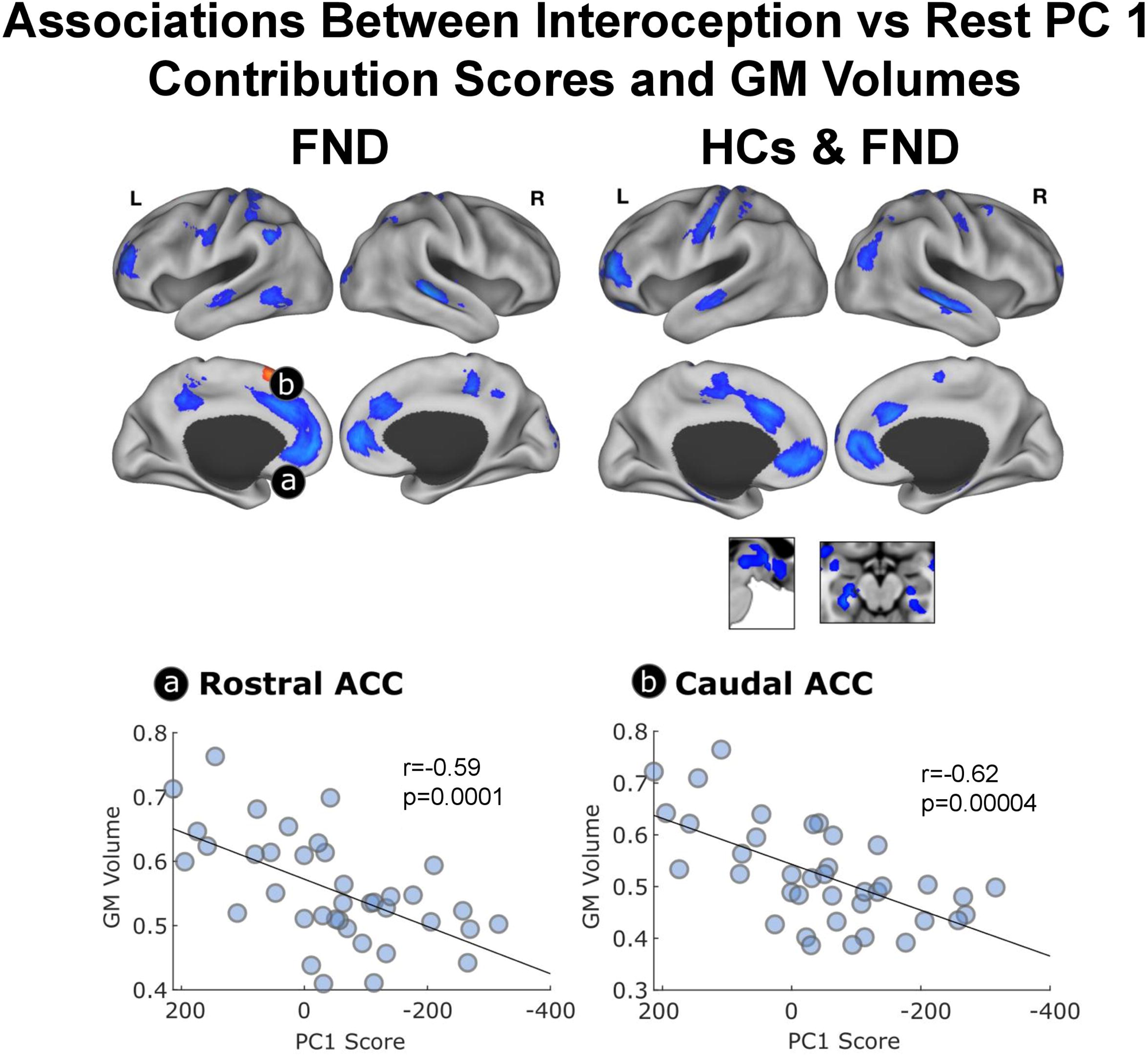
Within-group correlations between the grey matter (GM) volume and principal component (PC) 1 scores for interoception vs. rest contrast in patients with functional neurological disorder (FND) alone or FND and healthy control (HC) cohorts combined. Within the FND cohort, GM volume primarily in the ventral and dorsal prefrontal cortices – including the rostral and dorsal anterior cingulate cortex – correlated with the PC1 scores, suggesting that FND individuals with lower GM volume in these brain regions exhibited relatively attenuated interoception-related co-activations during the task. Across both FND and HC cohorts, GM volumes primarily in the ventral and dorsal prefrontal cortices – including the rostral and dorsal anterior cingulate cortex, medial temporal cortices, hippocampus and hypothalamus correlated with PC1 contribution scores.

### 3.4. Associations Between Principal Component 1 Scores, Cardiac Interoception, and Psychopathology Scores

In patients with FND, increasingly negative contribution scores to PC1 in the interoception vs. rest contrast positively correlated with increased dissociation, depression and anxiety scores (see **Figure 5**). Neither HACC nor ITPE scores showed statistically significant correlations with individual differences in PC1 contribution scores for the interoception vs. rest contrast in patients with FND. In the interoception vs. exteroception contrast, there were no statistically significant correlations between PC1 contribution scores and HACC, ITPE, DES, BDI-II and STAI-total scores in patients with FND (see **Supplementary Figure 2**).

**Figure 5.**
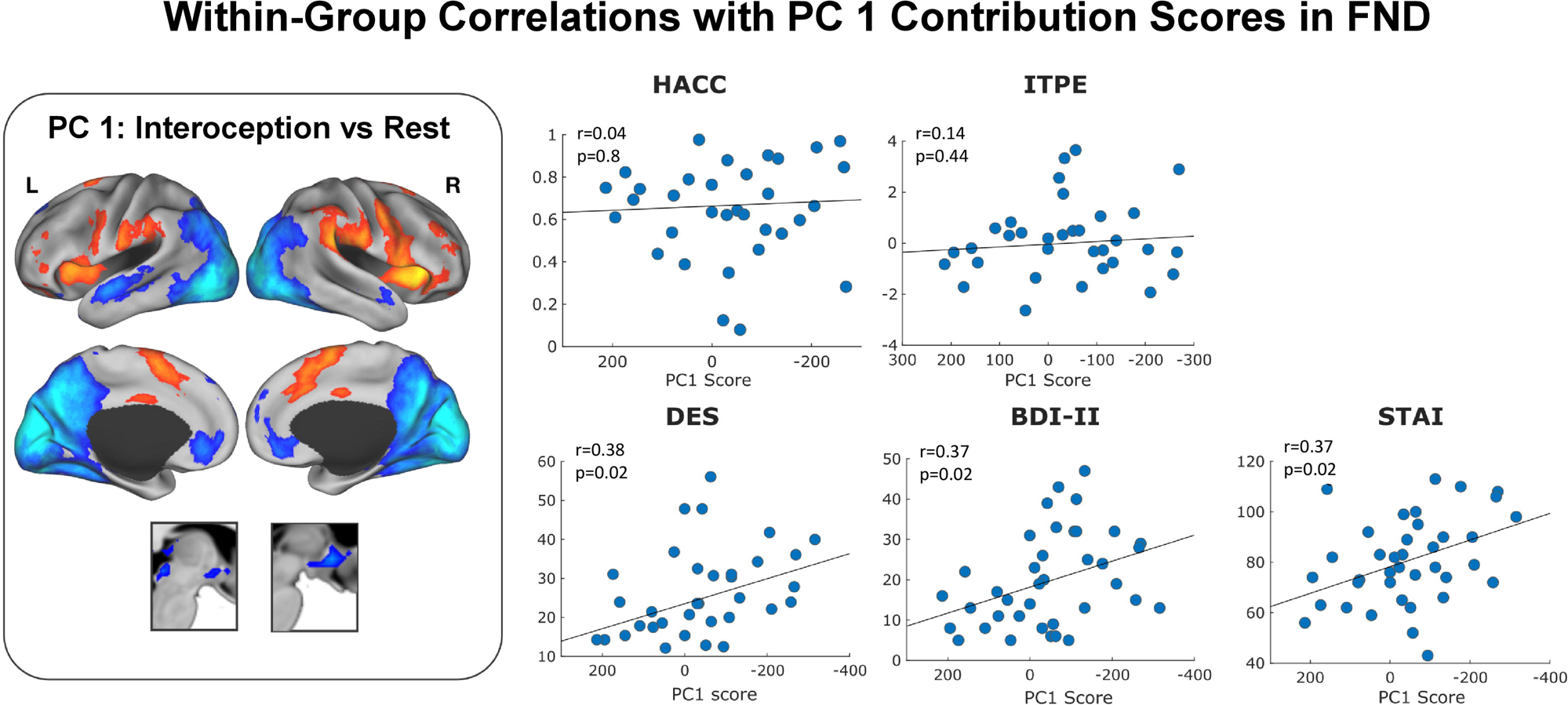
FND within-group correlations of the principal component (PC) 1 scores during interoception vs. rest with interoceptive and clinical variables. Across FND patients, dissociation, depression and anxiety scores correlated with increasingly negative PC1 contribution scores, indicating that higher symptom severity is associated with relatively attenuated co-activations in regions implicated in cardiac interoceptive processing vs. rest. Neither cardiac interoceptive accuracy nor ITPE related to PC1 contribution scores. HACC indicates heartbeat counting accuracy; ITPE, interoceptive trait prediction error; DES, Dissociation Experiences Scale-II; BDI-II, Beck Depression Inventory-II; STAI, Spielberger State Trait Anxiety Inventory.

In HCs, there were no significant correlations between PC1 contribution scores (in neither interoception vs. rest and interoception vs. exteroception contrasts) and HACC, ITPE and psychopathology scores.

## 4. Discussion

In this study, we combined a state-based fMRI assessment of cardiac interoception with VBM, self-report interoceptive measures, and dimensional psychopathology scores to examine multilevel profiles linked to interoception in FND. As expected, the heartbeat counting task increased salience and ventral attention network co-activations (e.g., bilateral anterior insula, middle cingulate gyri, and TPJ) across all participants as shown in PC1 of the interoception vs. rest and interoception vs. exteroception contrasts (Haruki & Ogawa, 2023; Schulz, 2016). Concurrently, we observed co-deactivations in default mode and visual processing networks in PC1, including ventromedial, posterior medial and occipital areas. In between-group analyses examining differences in PC contribution scores during cardiac interoceptive processing (vs. either rest or exteroception), the FND cohort compared to HCs in PC1 exhibited relatively attenuated co-activations in salience and ventral attentional networks, along with relatively less default mode and visual processing network co-deactivations. While individual differences in the PC1 contribution scores for the interoception vs. rest contrast did not relate to interoceptive accuracy or ITPE scores, PC1 contribution scores did correlate with dimensional psychopathology scores in patients with FND.

A noteworthy finding is the observation that patients with FND vs. HCs showed relatively blunted co-activations in bilateral anterior insular and middle cingulate cortices during cardiac interoceptive processing compared to both rest and exteroception. These findings provide novel evidence for attenuated activity of salience network cortical hubs during interoceptive processing in FND. Computational models of interoception highlight the salience network’s role in estimating precision of incoming sensory signals (including the importance/lack of importance of prediction errors) (Kleckner et al., 2017) consistent with accounts of salience network involvement in attentional regulation, multisensory integration and autonomic control of bodily homeostasis (Seeley, 2019). We speculate that the relatively attenuated cingulo-insular activation specific to interoception may reflect a disruption of precision-weighting of interoceptive signals in FND, leading to an ambiguous representation of afferent bodily information that is prone to aberrant categorization. Moreover, convergent meta-analytic and lesional evidence support the view that cingulo-insular regions are critical for interoceptive awareness – an important building block of affective experiences (Adolfi et al., 2017). Disrupted interoceptive signaling may therefore contribute to chronic pain, fatigue (Věchetová et al., 2018), dissociation (Koreki et al., 2020), alexithymia (Demartini et al., 2014) and autonomic dysregulation in patients with FND (Keynejad et al., 2019). Here, this is supported by increased anxiety, depression and dissociation severity scores associated with relative decreases in salience network co-activations in our FND cohort. Our findings are also consistent with abnormal cingulo-insular activations previously identified across the spectrum of FND during affective (Aybek et al., 2015; Goodman et al., 2022) and sensorimotor tasks, (Nahab et al., 2017; Voon et al, 2011) including our prior work (in a distinct FND cohort) where we identified correlations between alexithymia and decreased insular activation during viewing of unpleasant stimuli (Sojka et al., 2019).

Relatively blunted co-activations in the right TPJ were observed in patients with FND vs. HCs while attending to cardiac interoceptive signals - consistent with perceptual and attentional deficits reported in FND cohorts (Drane et al., 2020). In the FND literature, abnormal right TPJ activation has been interpreted as a failure to predict action consequences leading to impaired sense of self-agency (Maurer et al., 2016; Nahab et al., 2017; Voon et al., 2010). However, the role of right TPJ is likely context- and network-dependent given its ubiquitous involvement across different cognitive domains (Masina et al., 2022). The right anterior TPJ is part of both salience and ventral attentional networks, and has been implicated in attentional reorienting, tonic alertness, and salience detection in the context of attentional tasks (Masina et al., 2022). Moreover, the right TPJ is a prominent source of the P300, an ERP component associated with contextual updating of internal models, with increased amplitude following salient unexpected stimuli (Valakos et al., 2020). Reduced or absent P300 signals have been reported in patients with FND during stimulation of affected body parts suggesting abnormal body-related attention (Lorenz et al., 1998). We speculate that the relatively attenuated right TPJ co-activations as part of PC1 found in our FND cohort may relate to a failure to orient attention to interoceptive input.

Compared to HCs, patients with FND also displayed relatively less co-deactivations in bilateral vmPFC/anterior cingulate cortex, posterior cingulate cortex and precuneus during interoceptive processing. Moreover, lower GM volume in these regions in patients with FND correlated with attenuated recruitment of salience network areas during cardiac interoception vs. rest. The vmPFC is a core part of the default mode network, and is anatomically well positioned to integrate conceptual thought with peripheral physiology given its descending projections to autonomic and neuroendocrine control regions (e.g., hypothalamus and periaqueductal grey) (Koban et al., 2021). In our study, the hypothalamus and periaqueductal grey exhibited less co-deactivations during the heartbeat counting task vs. rest in the FND cohort compared to the HCs (Aybek et al., 2015). vmPFC hyperactivation has been previously associated with disruptions of bodily homeostasis (Jarrahi et al., 2015; Teves et al., 2004)and implicated across psychiatric disorders during processing of emotional stimuli (McTeague et al., 2020). Abnormal vmPFC activations have been observed in FND cohorts across a spectrum of motor tasks (Bègue et al., 2018; de Lange et al., 2010), suggesting that the abnormal modulation of sensorimotor processes may be associated with altered neural activity relevant for self-representations (Bègue et al., 2018). Posterior medial cortices including the posterior cingulate and precuneus form default mode network areas associated with self-referential cognition, mediation between internal and external focus (Leech & Sharp, 2014), and integration of emotional states and memory (Cavanna & Trimble, 2006)among other functions. Prior FND studies showed abnormal posterior cingulate cortex/precuneus activations in response to negative emotional stimuli (Sojka et al., 2019), and altered precuneus connectivity and cortical thickness related to FND symptom severity and illness duration (Mueller et al., 2022; Zelinski et al., 2022). Computational models of homeostatic and allostatic regulation propose that these default mode network hubs participate in body regulation by adaptively assigning personal meaning and value to interoceptive afferents with respect to past experience (Smith et al., 2017). Collectively, the observations of relatively blunted salience network co-activations and relatively less default mode network co-deactivations during cardiac interoceptive processing in patients with FND vs. HCs suggest that the co-activation balance between networks may be important considerations in the pathophysiology of FND.

Additionally, FND patients co-deactivated the visual processing network less than HCs during interoceptive processing. Greater occipital co-deactivations for HCs during interoception relative to exteroception is consistent with previous EEG studies reporting suppression of parieto-occipital activity during internally-oriented attention (Vilena-González et al., 2017). While we cannot exclude other possibilities, we speculate that the relatively higher occipital activations in patients with FND may be associated with reduced attentional allocation to interoceptive signals. However, recent studies also reported occipital regions activations in interoception that require further exploration (Haruki & Oqawa, 2023; Wang et al., 2019).

Limitations of this study include the modest sample size, psychotropic medication use, and phenotypic heterogeneity. We adopted a transdiagnostic approach across FND-seiz and FND-movt given arguments by several groups that these populations are on a clinical continuum (Perez et al., 2021). Since this is still debated, a strength of our results is that between- and within-group findings held adjusting for FND subtype, supporting a neurobiological overlap between FND-seiz and FND-movt populations. Using only healthy subjects as the comparison group limits our ability to verify the specificity of observed findings to FND; future research should also incorporate psychiatric and/or neurological control groups. While we utilized staircase thresholding of tone stimuli to try to match the difficulty of exteroceptive and interoceptive conditions, both HCs and patients were more accurate in tone counting; thus, we cannot exclude the influence of uneven condition difficulty on the present findings. Given that events precipitating FND onset commonly involve bodily (homeostatic) perturbations, future studies should consider experimentally modulating interoceptive signals and exploring dynamic changes in interoception on FND symptomatology. Moreover, given that non-motor symptoms have been identified as primary drivers of health-related quality of life in FND patients (LaFrance & Syc, 2009; Věchetová et al., 2018),future research should specifically aim to characterize relationships between interoceptive accuracy, non-motor symptoms and neural circuit activation profiles. Additionally, our findings suggest that interventions using interoceptive awareness approaches (biofeedback, mindfulness-based therapy etc.) require more research in FND.

In conclusion, this multimodal neuroimaging study identified attenuated salience network co-activations and blunted default mode network co-deactivations during interoceptive processing in a cohort of FND patients relative to HCs. Individual differences in dimensional psychopathology scores mapped onto these co-activation/co-deactivation patterns. This study provides important pathophysiology insights across the spectrum of FND-seiz and FND-movt, underscoring the need for additional research on interoception in this population.

---

**Supplementary Figure 1. Within-group correlations between the grey matter (GM) volume and principal component (PC) 1 scores for interoception vs. exteroception contrast in patients with functional neurological disorder (FND) alone or FND and healthy control (HC) cohorts combined.**

**Supplementary Figure 2. Within-group correlations of principal component (PC) 1 contribution scores computed from the interoception > exteroception contrast with interoceptive and clinical variables in functional neurological disorder patients.** HACC indicates heartbeat counting accuracy; ITPE, interoceptive trait prediction error; DES, Dissociation Experiences Scale-II; BDI-II, Beck Depression Inventory-II; STAI, Spielberger Anxiety Inventory-State.

## Funding

This study was supported by the Czech Ministry of Health Project AZV NU20-04-0332 and NW24-04-00456 (P.S. and T.S.); National Institute for Neurological Research project (Programme EXCELES, ID Project No. LX22NPO5107) - Funded by the European Union – Next Generation EU (P.S. and T.S.); Cooperatio Program in Neuroscience at Charles University (P.S. and T.S.).

## Conflicts of Interests / Disclosures

P. Sojka reports no disclosures relevant to the manuscript; T. Serranová reports no disclosures relevant to the manuscript; S.S. Khalsa is on the editorial board of *Biological Psychology*, has performed scientific consultation for Janssen, is supported by funding from the NIH and The William K. Warren Foundation, and has no conflicts of interest or disclosures related to this work; D.L. Perez has received royalties from Springer Nature for a functional movement disorder textbook, honoraria for continuing education lectures on functional neurological disorder and for an Elsevier textbook on functional neurological disorder, is a paid editor of *Brain and Behavior*, is on the editorial board of *Epilepsy & Behavior*, *The Journal of Journal of Neuropsychiatry & Clinical Neurosciences*, and *Cognitive and Behavioral Neurology*, and has received funding from the NIH and Sidney R. Baer Jr. Foundation unrelated to this work; I. Diez reports no disclosures relevant to the manuscript.

## Acknowledgements

Multimodal and Functional Imaging Laboratory of CEITEC Masaryk University is gratefully acknowledged for the obtaining of the scientific data presented in this study.

## Contributions

Petr Sojka: Drafting/revision of the manuscript for content, including medical writing for content; Major role in the acquisition of data; Study concept or design; Analysis or interpretation of data.

Tereza Serranová: Drafting/revision of the manuscript for content, including medical writing for content.

Sahib S. Khalsa: Drafting/revision of the manuscript for content, including medical writing for content.

David L. Perez: Drafting/revision of the manuscript for content, including medical writing for content; Analysis or interpretation of data.

Ibai Diez: Drafting/revision of the manuscript for content, including medical writing for content; Analysis or interpretation of data.

## Notes

### Competing Interest Statement

P. Sojka reports no disclosures relevant to the manuscript; T. Serranova reports no disclosures relevant to the manuscript; S.S. Khalsa is on the editorial board of Biological Psychology, has performed scientific consultation for Janssen, is supported by funding from the NIH and The William K. Warren Foundation, and has no conflicts of interest or disclosures related to this work; D.L. Perez has received royalties from Springer Nature for a functional movement disorder textbook, honoraria for continuing education lectures on functional neurological disorder and for an Elsevier textbook on functional neurological disorder, is a paid editor of Brain and Behavior, is on the editorial board of Epilepsy & Behavior, The Journal of Journal of Neuropsychiatry & Clinical Neurosciences, and Cognitive and Behavioral Neurology, and has received funding from the NIH and Sidney R. Baer Jr. Foundation unrelated to this work; I. Diez reports no disclosures relevant to the manuscript.

### Funding Statement

This study was supported by the Czech Ministry of Health Project AZV NU20-04-0332 (P.S. and T.S.); National Institute for Neurological Research project (Programme EXCELES, ID Project No. LX22NPO5107), Funded by the European Union, Next Generation EU (P.S. and T.S.); Cooperatio Program in Neuroscience at Charles University (P.S. and T.S.).

### Author Declarations

Ethics committee of Masaryk University (Brno, Czech Republic) and St. Anne's University Hospital (Brno, Czech Republic) gave ethical approval for this work

