## Supplementary figures and images for "Interoceptive disruption in functional neurological disorder: a multimodal brain imaging study"

### Supplementary Figure 1

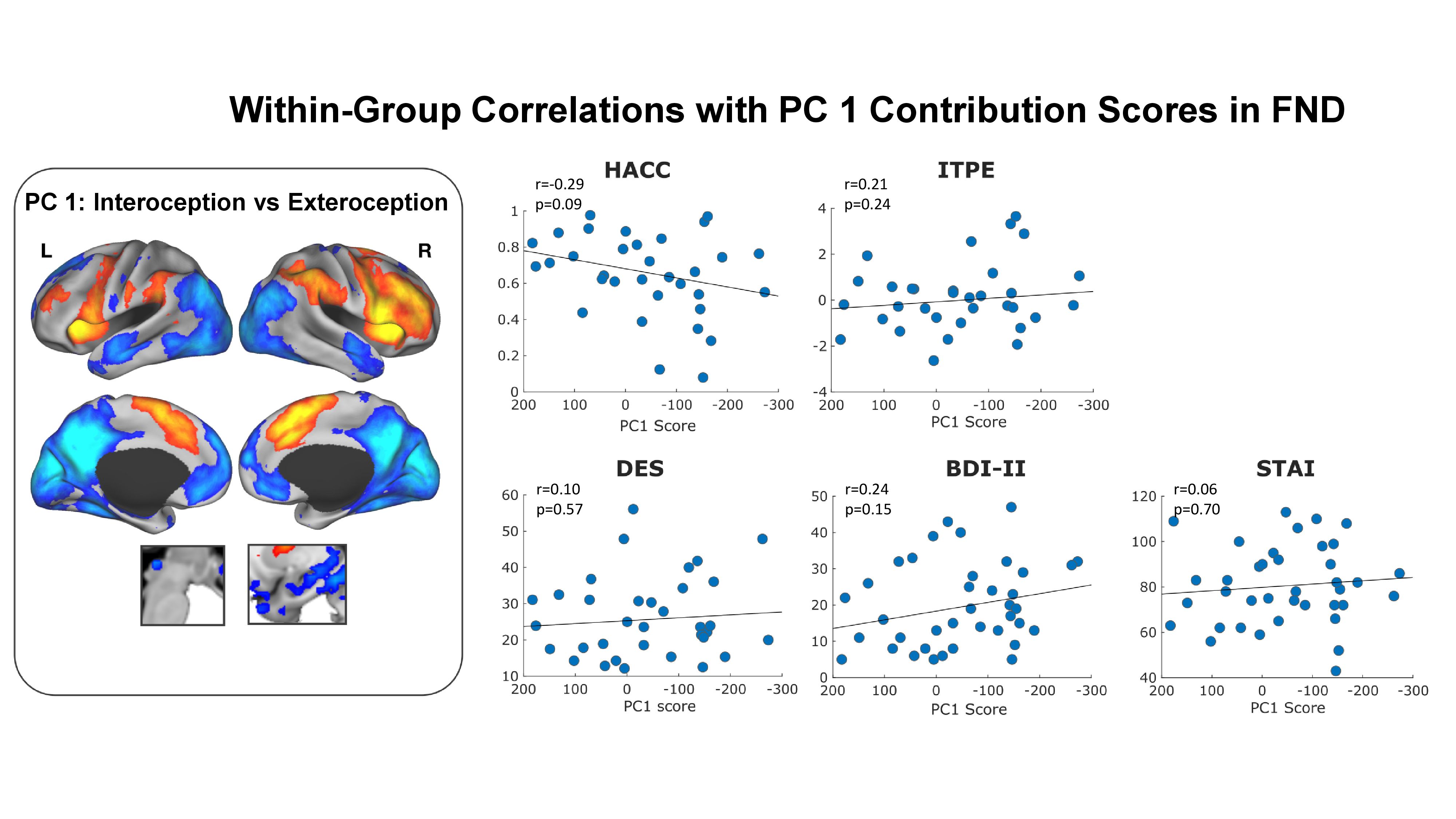

### Supplementary Figure 2

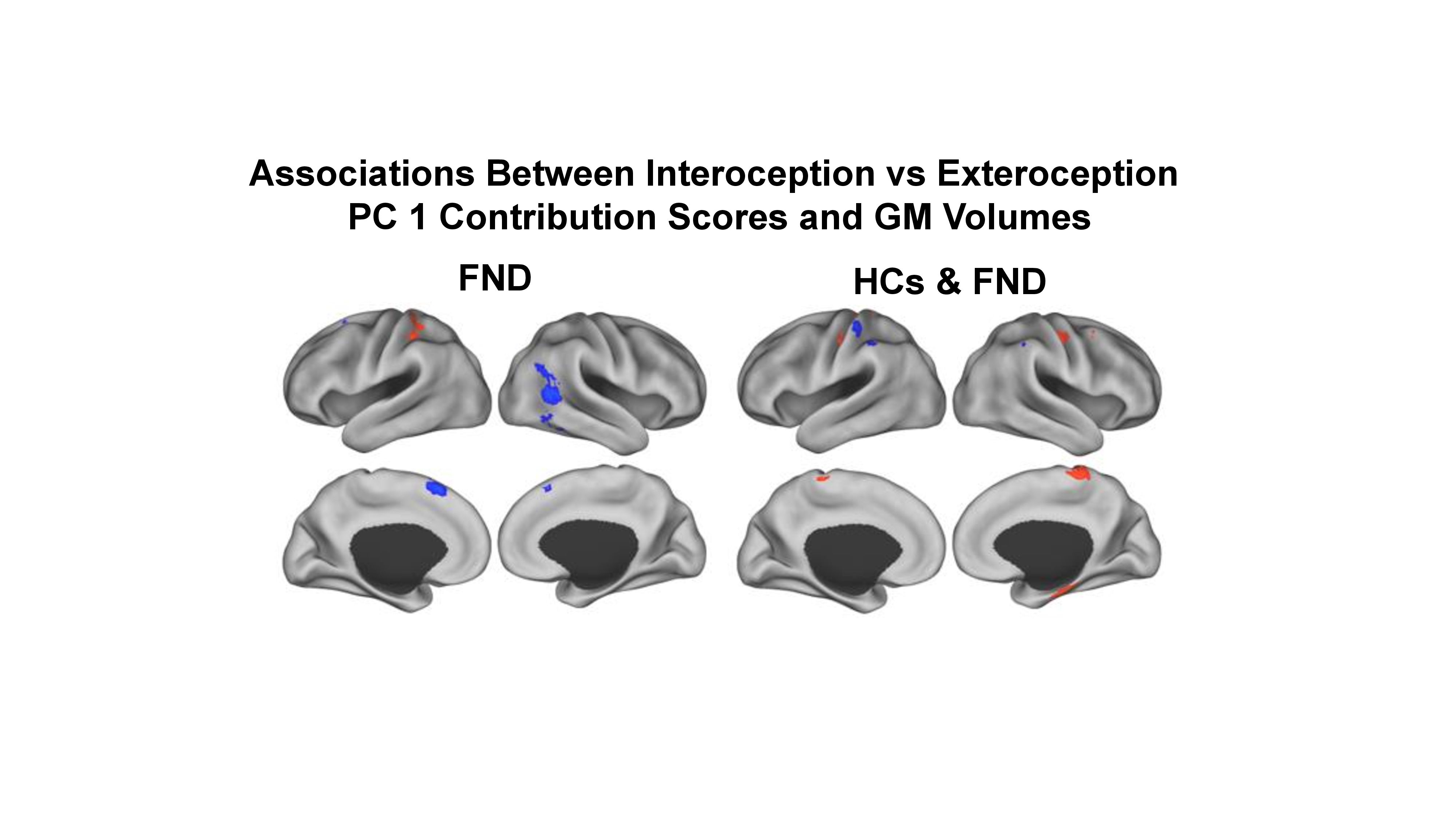
